# Testing new versions of ChatGPT in terms of physiology and electrophysiology of hearing: improved accuracy but not consistency

**DOI:** 10.1101/2024.10.08.24315089

**Authors:** W. Wiktor Jędrzejczak, Henryk Skarżyński, Krzysztof Kochanek

## Abstract

**Introduction:** ChatGPT has revolutionized many aspects of modern life, including scientific ones. Since its introduction, new versions have been introduced and advertised as having better performance. But is this true? This study aimed to assess the accuracy and consistency of six versions of ChatGPT (3.5, 4, 4o mini, 4o, 4o1 mini, and 4o1 preview). Of interest was the variability of responses given to asking the same question multiple times.

**Methods:** We evaluated 6 versions of ChatGPT based on their responses to 30 single-answer, multiple-choice exam questions from a 1-year course on objective methods of testing hearing. The questions were posed 10 times to each version of ChatGPT across two days (5 times each day). The accuracy of the responses was evaluated in terms of a response key. To evaluate consistency (repeatability) of the responses over time, percent agreement and Cohen’s Kappa were calculated.

**Results:** The overall accuracy of ChatGPT increased with each version, starting from around 53% for version 3.5 and rising to 86% for version 4o1 preview. The greatest improvement in accuracy and repeatability came with the introduction of version 4o. Repeatability progressively rose with newer releases with the exception of version 4o1 mini. While the current top version 4o1 preview has similar repeatability to 4o, the faster version, 4o1 mini, had significantly lower repeatability than the older 4o mini.

**Conclusion:** Newer versions of ChatGPT generally show improvement in terms of accuracy, but not in repeatability. The variability of responses is probably the current main limitation of ChatGPT for professional applications. Users must be especially careful with version 4o1 mini.

## Introduction

Artificial intelligence (AI) systems based on large language models (LLMs) have undergone significant advances in recent years, profoundly impacting various aspects of society, including science, education, healthcare, and industry [1–3]. Among LLMs, OpenAI’s ChatGPT has emerged as a prime contender. For example, when PubMed was searched for “chatgpt” it yielded 2774 papers for 2024 alone (search performed on 01.10.2024) while LLMs from other developers yielded far fewer. Within many medical fields, ChatGPT far outnumbers other LLMs [4,5]. Since its initial release, many versions of ChatGPT have been developed, and with each release the manufacturer claims improved overall performance [6].

In 2024 OpenAI released four new versions of ChatGPT. Until May 2024 there had been two versions (3.5 released in November 2022 and 4 in March 2023), with 4 claiming better performance [7,8]. In May of this year versions 4o mini and 4o were released, with 4o mini being faster while 4o claimed better results [6]. At that time, version 3.5 was discontinued. In September 2024 updates were made: 4o1 mini and 4o1 preview (with the former faster and the latter providing a full range of features) [9]. Version 4o1 preview is advertised as spending more time thinking but excelling in math and coding [9]. The newer versions open up new horizons but may also meet unexpected challenges. While enhancements are expected at each iteration, perhaps the speed of development increases the risk of mistakes.

In a professional or academic setting, accuracy and consistency are critical factors for the acceptability of an AI language model [8,10,11]. In a field such as in medical education or scientific research, precision is essential and so any mistakes will undermine trust in AI. At the same time, different responses to the same question will not be acceptable.

The motivation for this study is that there are presently only a small number of studies that have systematically evaluated the comparative performance of different versions of ChatGPT. So far, studies have shown that there has been an improvement in performance from ChatGPT 3.5 to 4, and even better to 4o [8,12–15]. At the time of writing there seems to have been no evaluation of version 4o mini (Pubmed search “ChatGPT 4o mini” yield 0 results at 01.10.2024) or evaluations of versions 4o1 in the scientific or medical sphere. This gap prevents practitioners and researchers from gauging progress in the field and choose the best version for their needs.

Here we focus on the field of audiology and in particular on topics related to the physiology and electrophysiology of hearing. Audiology seems underrepresented compared to other fields in which ChatGPT has been tested, with only a few studies available, e.g. [4,8,16,17]. Audiology is a specialized domain requiring specific knowledge, and so provides a rigorous test of LLM abilities.

The present study therefore aims to quantitatively assess the accuracy and consistency of six versions of ChatGPT: 3.5, 4, 4o mini, 4o, 4o1 mini, and 4o1 preview and assess what progress has been made. We employed a set of single-answer, multiple-choice exam questions from a one-year course on objective methods of testing hearing. The established answers were used as the yardstick for testing the correctness and consistency of the answers given by ChatGPT when the same question was asked many times.

## Methods

The responses of six versions of OpenAI’s chatbot ChatGPT to a set of single-choice questions were evaluated. The versions tested were ChatGPT 3.5, 4, 4o mini, 4o, 4o1 mini, and 4o1 preview (in order of less to more advanced). They were made public in November 2022 (3.5), March 2023 (4), May 2024 (4o mini and 4o), and September 2024 (4o1 mini and 4o1 preview). The questions related to physiological measurements of hearing status, in particular tympanometry, middle ear muscle responses, otoacoustic emissions, and auditory brainstem responses. The questions are the same as used in exam questions for a one-year course in objective methods of testing hearing. The questions and response key are provided as an appendix to a previous paper which evaluated ChatGPT 3.5 and 4 [8] and they are also attached here as supplementary material.

In general the approach here is quite similar to before but considers more versions of ChatGPT [8]. The questions were presented to each version of ChatGPT 10 times (5 times one day, and 5 times the next) to take into account some variability in responses. The variability of ChatGPT can be changed to some extent when using it via the application programming interface (API) [18–20]. It is controlled by the temperature parameter, which can be set to values from 0 to 2 (from lower variability to higher). The default setting is 1 and the ChatGPT application runs on this setting. We decided to use ChatGPT in default mode to be as close as possible to the typical user. This is also further explained in the discussion section.

All versions except 3.5 were tested in September 2024. Version 3.5 was then not available. However we did perform the same test at the end of April 2024 shortly before this version was shutdown (May 2024). To check whether there had been no changes in performance during time we compared the results of version 4 given in April 2024 and in September 2024 and found no significant differences. This seems to justify the inclusion of a comparison of version 3.5 with all the other versions.

All analyses were made in Matlab (version 2023b, MathWorks, Natick, MA). Percent agreement was used to evaluate correctness and consistency (test–retest repeatability) of the responses. By correctness of responses the percent of correct responses versus response key was calculated. For consistency the correctness was ignored. Instead, each one of the 10 trials was compared with the other 9. If the responses were always the same we gave a rating of 100% repeatability (even if the response was incorrect). Additionally, Cohen’s Kappa [19] was used to rate test–retest repeatability. As the data had a non-normal distribution, a Friedman test was used to assess the effect of using different versions. For pairwise comparisons a nonparametric Mann–Whitney 𝔘 -test was used. In all analyses, a 95% confidence level (p < 0.05) was taken as the criterion of significance. Multiple comparisons were corrected using the Benjamini–Hochberg procedure [21].

## Results

The average accuracy of responses (i.e. correctness based on the response key) for all six versions of ChatGPT is shown in Figure 1. It ranged from around 53% for version 3.5 to 86% for version 4o1 preview. The more broadly available (free of charge) versions, 4o mini and 4o1 mini, achieved 63% and 75% respectively. Average accuracy for all versions was compared using a Friedman test and revealed significant differences (Chi-sq = 46.5, *p* < 0.001). Pairwise comparisons revealed statistically significant differences between all versions except 4 and 4o mini. It can be seen that there was a general improvement with time. Also the higher priced top models (4, 4o, and 4o1 preview) generated better overall results than the base versions (3.5, 4o mini, 4o1 mini). The figure also indicates the most frequent responses out of ten trials. It can be seen that they differ slightly from the average values.

**Figure 1.**
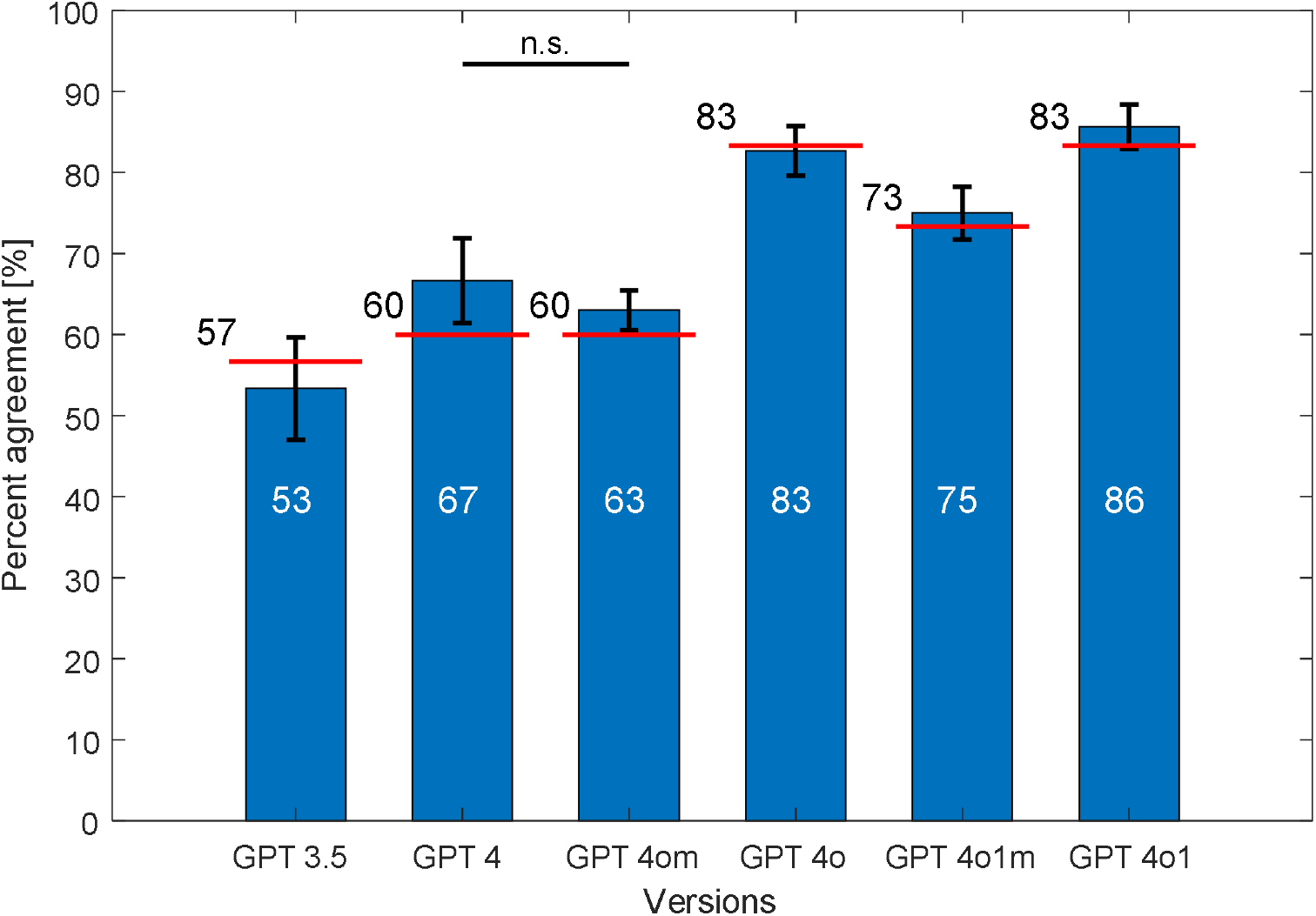
Average percent agreement of six versions of ChatGPT in response to a set of multiple choice questions. ChatGPT versions are shown from older (left) to newer (right). The average percentage score is shown on each bar in white. The red lines indicate the percentage agreement of the most frequent responses in different samples (percentage given next to the line). Whiskers indicate standard deviations. All differences are statistically significant except that marked n.s.

An example of a mistake can be seen in response to question 3: “The frequency of the measuring tone for tympanometry in a child aged 3 months should be:”, which was often given as 220 Hz or 226 Hz for versions 3.5, 4, 4o mini, and 4o1 mini. In current clinical practice the correct answer is 1000 Hz, a frequency recommended for children older than 3 months [22,23]. The question isn’t tricky, since the debate over 226 Hz and 200 Hz was settled long ago, and this error seems surprising. One good feature is that it was answered correctly by the top versions, 4o and 4o1 preview.

The next step was to evaluate the percentage of questions receiving a correct answer on all 10 trials for each version of ChatGPT, and the results are shown in Figure 2. For ChatGPT 3.5, the fraction of questions that were answered correctly on all 10 trials was 40%, but was higher for all the other versions. Versions 4, 4o mini, and 4o1 mini exceeded 50% but were less than 60%. At the top were versions 4o and 4o1 preview, which both attained 77%. An especially interesting aspect was the variability of responses, which we expected might decrease with higher versions, but the picture wasn’t clear. The fraction of variable responses remained constant at 30% for versions 3.5, 4, and 4o1 mini. It was higher for 4o1 preview (16.7%) than the earlier 4o (10%), although the fraction of wrong responses was lower for 4o1 preview.

**Figure 2.**
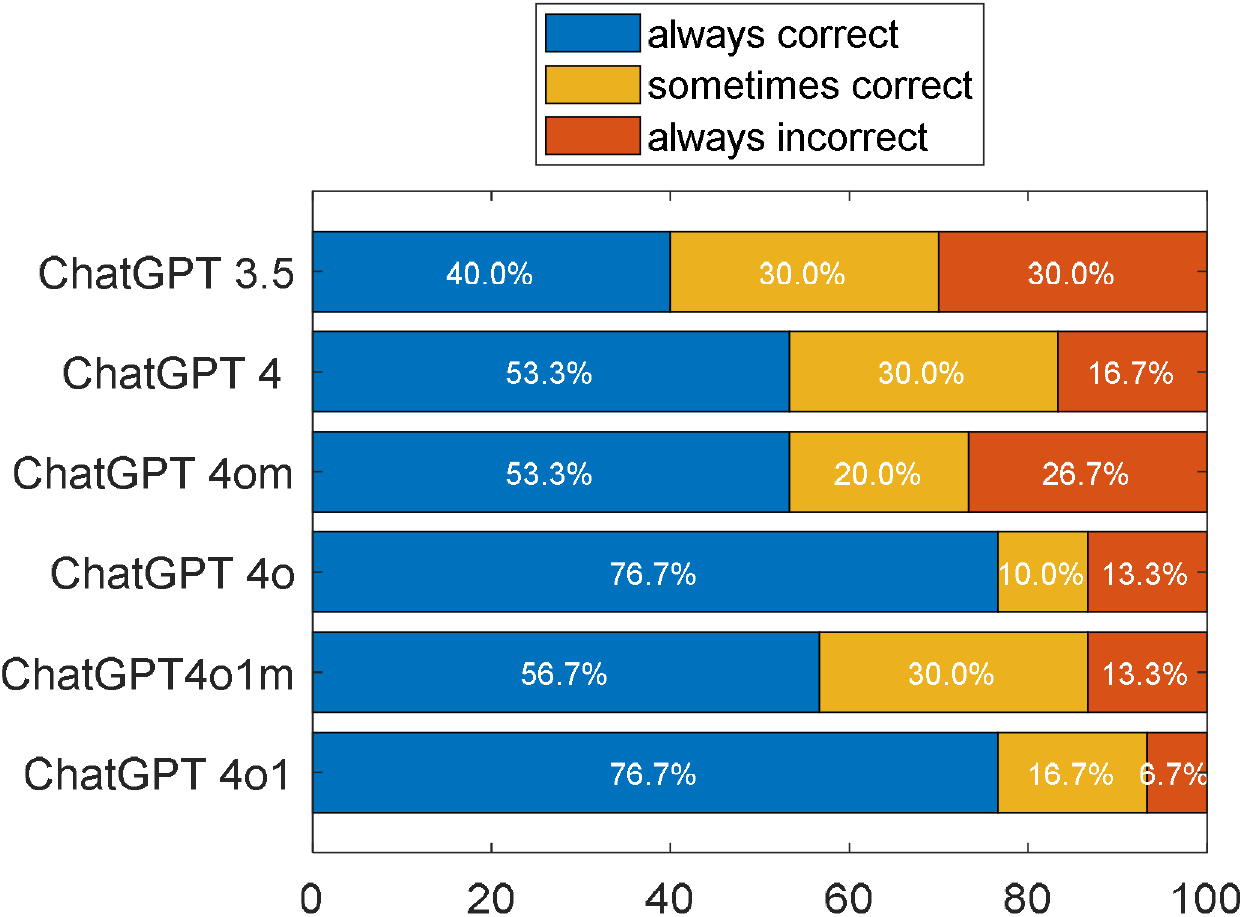
Six versions of ChatGPT and their correctness in answering 30 questions across 10 trials. The colours show percentage of questions that were always answered correctly (blue), sometimes correctly and sometimes incorrectly (yellow), and always incorrectly (red).

We explored this aspect further by focusing solely on repeatability, i.e. comparing the responses between trials without taking into account correctness, and the findings are shown in Figure 3. Average repeatability for all versions was compared by Friedman test and revealed significant differences (Chi-sq = 153.0, *p*< 0.001). Pairwise comparisons revealed statistically significant differences between most versions (the exceptions being 4 and 4o mini, 4 and 4o1 mini, and 4o and 4o1 preview). While the repeatability generally increased from 3.5 to 4o, it then dropped for 4o1 mini. It appears that 4o1 mini has significantly higher repeatability only from 3.5 onwards but has lower repeatability than the previous 4, 4o mini, and 4o versions. Table 1 compares repeatability in terms of Cohen’s Kappa, where values range from 0.71 for version 3.5 to 0.92 for 4o. For all versions Cohen’s Kappa was significant (*p* < 0.001) which means that agreement between trials did not occur by chance.

**Table 1.** Consistency between responses. In all cases Cohen’s Kappa had p<0.001, indicating that observed agreement was not accidental.

|  | Cohen's Kappa |  |
| --- | --- | --- |
| ChatGPT 3.5 | 0,71 | 0,07 |
| ChatGPT 4 | 0,78 | 0,06 |
| ChatGPT 4o mini | 0,80 | 0,07 |
| ChatGPT 4o | 0,92 | 0,04 |
| ChatGPT 4o1 mini | 0,75 | 0,07 |
| ChatGPT 4o1 | 0,91 | 0,05 |

**Figure 3.**
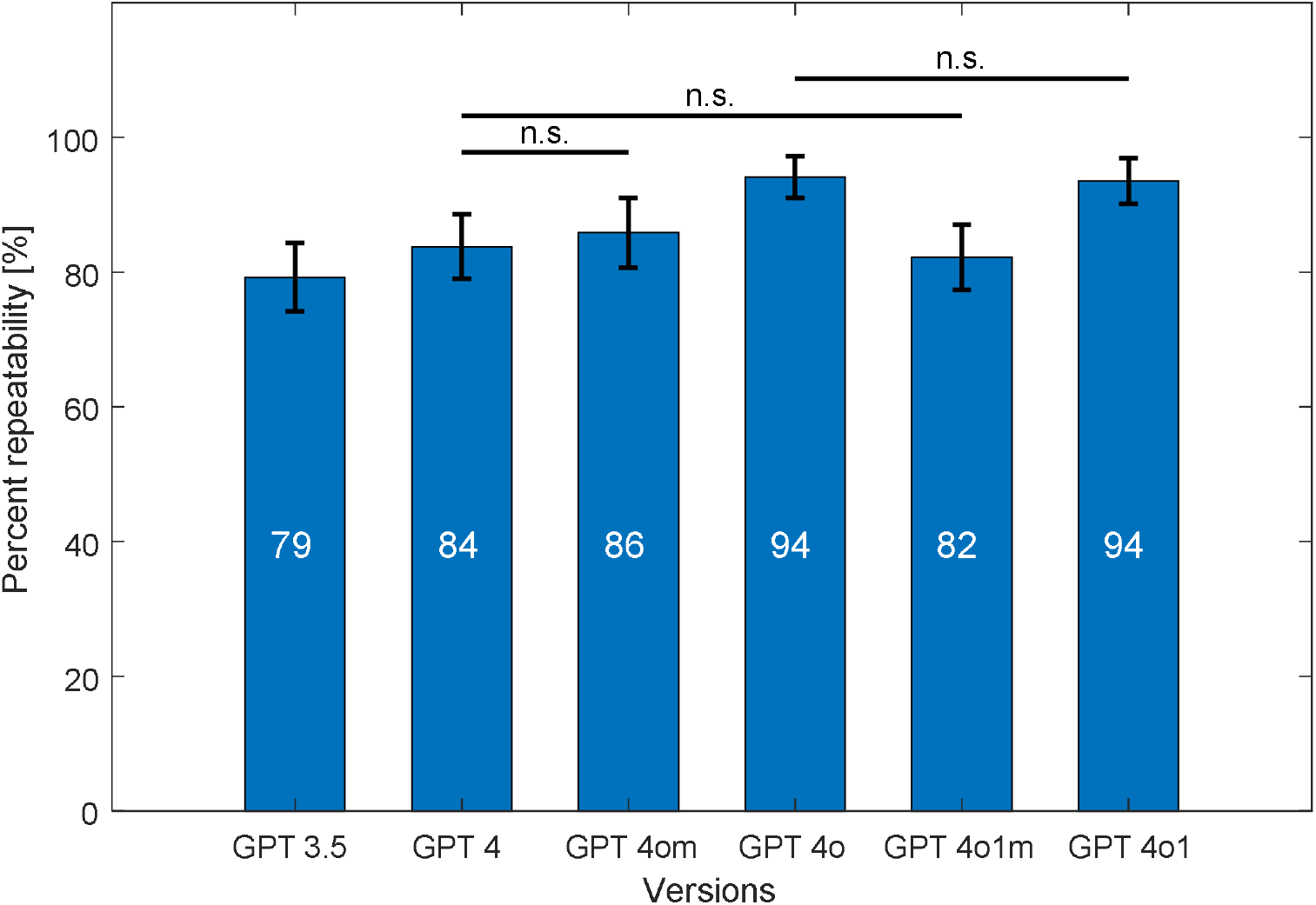
Six versions of ChatGPT (older to newer) and their average percent repeatability in response to a set of multiple choice questions. The average percentage score is shown on each bar in white. Whiskers are standard deviations. All differences are statistically significant except when marked n.s.

## Discussion

Four new versions of ChatGPT were released this year. To our knowledge, this study represents one of the first evaluations in which the versions are compared. We set out to investigate if newer versions of ChatGPT really have improved performance. The results show that the issue is not clear-cut. While later versions are generally better, they have shortcomings in terms of consistency.

Our evaluation revealed a clear trend of improvement from version 3.5 to version 4o. Specifically, version 3.5 achieved an average accuracy of approximately 53%, while version 4o reached around 86%. This trend suggests that model updates have improved performance. However, the introduction of the o1 versions has been accompanied by some inconsistencies. The top-tier version, 4o1 preview, was better than its predecessors in accuracy, achieving 86% while maintaining consistency. In contrast, version 4o1 mini, despite an improvement in accuracy over 4o mini (from 63% to 75%), exhibited reduced consistency.

Currently, there are no published studies evaluating versions 4o1 preview and 4o1 mini, limiting direct comparisons with other work. However, previous research has shown steady improvements in earlier versions. For instance, some studies have reported improvements in accuracy from approximately 38–60% for version 3.5 to 52–81% for version 4 and 69–90% for 4o (e.g. [8,12,14,15]). These scores, while sometimes slightly worse or better than ours, generally align with our tests, reinforcing the notion of progressive enhancement in newer models.

Surprisingly, we did not find prior evaluations of the ‘mini’ versions. Given their greater accessibility and faster operation, it is plausible that these versions are more frequently used in everyday applications than their advanced counterparts like 4o and 4o1 preview. Our results suggest that while the mini versions are more accessible, they may compromise on consistency and accuracy, which are critical in a professional setting. Nevertheless, it should be said that their performance is on a level comparable with version 4, which was the top version only a few months ago.

In our opinion, consistency of responses is as important as accuracy. Of course, if generating dialogue or different personalities, variability may be an advantage [24–27], but uncontrolled variability in a specific discipline, especially medicine, may cause considerable harm [20]. For example, a patient confronted with two different answers may neglect to get treatment. It seems likely that variability of ChatGPT responses is connected to the phenomenon known as “hallucinations.” Hallucinations refer to instances where AI language models generate information that is incorrect or entirely fabricated while presenting it confidently as factual [28]. The occurrence of hallucinations may be influenced by factors such as training data quality, model architecture, imprecise posing of a question, and also innate variability [29]. The problem is amplified because ChatGPT usually fails to provide sources of information [16]. If asked for sources, it frequently makes them up. Unfortunately the result looks very professional and only careful verification can detect falsification [30,31]. Some errors are tiny, e.g. one digit different in the DOI of a paper [31].

It should be mentioned that the variability can be controlled to some extent by the temperature parameter [20], but this is only possible when using ChatGPT via the API. Different settings of this parameter can produce different results. It has been shown that changing this parameter to a lower variability setting did not make significant changes in the correctness of responses for ChatGPT version 4, but for version 3.5, it caused a decrease in correctness [13]. This shows little benefit to users trying to modify this setting. Therefore, in this study, we used the default setting for this parameter, as it seems to be the most commonly used and provides the best results [13]. Response variability can also be controlled to some extent by asking ChatGPT the same question multiple times and selecting the most common answer. In some cases, this can provide better results (see Figure 1). Nevertheless, it seems unlikely that a typical user looking for a simple answer would do this.

Another important issue is that improvements over time may be due not from model enhancements but from more extended training and updates. Continuous training improves performance, with many studies indicating that AI responses improve with ongoing use and data exposure [16]. We do not have detailed information about this process as it is an in-house operation. This is a general issue with AI development, and the great speed of development and lack of transparency has the potential to generate major errors.

The findings from this study have significant implications for both users and developers of AI based on LLMs. The fact that version 4o1 mini, despite higher accuracy, demonstrated lower consistency than 4o mini underscores the importance of balancing these two aspects. Users should be aware of these trade-offs when selecting a model for specific applications, particularly in fields where reliability is paramount. For developers, the results highlight the necessity of focusing development efforts not just on accuracy but also on enhancing response consistency.

On a positive note, the steady improvement in accuracy across versions is encouraging. Although none of the versions surpassed the 90% accuracy threshold in our test, the upward trend suggests that achieving such performance may be attainable in the near future. This progression bodes well for the integration of AI technologies into professional practice, provided that issues of consistency are addressed.

Ultimately, this paper wishes to prompt a broader conversation about the responsible and effective integration of AI technologies into professional domains. While the advances so far are promising, there remains substantial room for improvement. Future development should prioritize enhancing both accuracy and consistency to ensure that AI language models can serve as reliable tools in critical fields like healthcare.

## Conclusions

This study has demonstrated that newer versions of ChatGPT are better in terms of accuracy, but inconsistent responses persist. In a specialized area such as the physiology and electrophysiology of hearing, where precise and consistent information is crucial, inconsistencies pose challenges and potential risks. The same is true in medicine more generally.

The much improved performance of version 4o1 preview demonstrates that progress is being made. At the same time, the lower consistency of its counterpart, version 4o1 mini, underscores the gap that still exists in achieving both high accuracy and consistency.

For use in professional and clinical contexts, it is essential to exercise caution and verify all AI-generated information from authoritative sources. For developers, these findings emphasize the need to prioritize response consistency as much as accuracy in future iterations. Balancing these factors is vital for the safe and effective integration of AI large language models into specialized domains. Future efforts should focus on refining both reliability and consistency, thereby making them more suitable for use in professional practice.

## Data Availability

All data produced in the present study are available as a supplementary file.

## References

1. Nazi ZA, Peng W: Large Language Models in Healthcare and Medical Domain: A Review. Informatics. 2024, 11:57. 10.3390/informatics11030057

2. Birhane A, Kasirzadeh A, Leslie D, Wachter S: Science in the age of large language models. Nat Rev Phys. 2023, 5:277–80. 10.1038/s42254-023-00581-4

3. Suwała S, Szulc P, Guzowski C, et al.: ChatGPT-3.5 passes Poland’s medical final examination-Is it possible for ChatGPT to become a doctor in Poland? SAGE Open Med. 2024, 12:20503121241257777. 10.1177/20503121241257777

4. Jedrzejczak WW, Kochanek K: Comparison of the Audiological Knowledge of Three Chatbots: ChatGPT, Bing Chat, and Bard. Audiol Neurootol. 2024, 1–7. 10.1159/000538983

5. Fabijan A, Zawadzka-Fabijan A, Fabijan R, Zakrzewski K, Nowosławska E, Polis B: Assessing the Accuracy of Artificial Intelligence Models in Scoliosis Classification and Suggested Therapeutic Approaches. J Clin Med. 2024, 13:4013. 10.3390/jcm13144013

6. Introducing GPT-4o and more tools to ChatGPT free users. Accessed: October 5, 2024. https://openai.com/index/gpt-4o-and-more-tools-to-chatgpt-free/.

7. N O, Gs C, Wy L: ChatGPT goes to the operating room: evaluating GPT-4 performance and its potential in surgical education and training in the era of large language models. Ann Surg Treat Res. 2023, 104:. 10.4174/astr.2023.104.5.269

8. Kochanek K, Skarzynski H, Jedrzejczak WW: Accuracy and Repeatability of ChatGPT Based on a Set of Multiple-Choice Questions on Objective Tests of Hearing. Cureus. Published Online First: 8 May 2024. 10.7759/cureus.59857

9. Introducing OpenAI o1. Accessed: October 5, 2024. https://openai.com/index/introducing-openai-o1-preview/.

10. Cavalcanti RB, Sibbald M: Am I Right When I Am Sure? Data Consistency Influences the Relationship Between Diagnostic Accuracy and Certainty. Acad Med. 2014, 89:107. 10.1097/ACM.0000000000000074

11. Franc JM, Cheng L, Hart A, Hata R, Hertelendy A: Repeatability, reproducibility, and diagnostic accuracy of a commercial large language model (ChatGPT) to perform emergency department triage using the Canadian triage and acuity scale. Can J Emerg Med. 2024, 26:40–6. 10.1007/s43678-023-00616-w

12. Liu C-L, Ho C-T, Wu T-C: Custom GPTs Enhancing Performance and Evidence Compared with GPT-3.5, GPT-4, and GPT-4o? A Study on the Emergency Medicine Specialist Examination. Healthcare. 2024, 12:1726. 10.3390/healthcare12171726

13. Rosoł M, Gąsior JS, Łaba J, Korzeniewski K, Młyńczak M: Evaluation of the performance of GPT-3.5 and GPT-4 on the Polish Medical Final Examination. Sci Rep. 2023, 13:. 10.1038/s41598-023-46995-z

14. Bicknell BT, Butler D, Whalen S, et al.: Critical Analysis of ChatGPT 4 Omni in USMLE Disciplines, Clinical Clerkships, and Clinical Skills. JMIR Med Educ. Published Online First: 14 September 2024. 10.2196/63430

15. Güneş YC, Cesur T, Çamur E, Günbey Karabekmez L: Evaluating text and visual diagnostic capabilities of large language models on questions related to the Breast Imaging Reporting and Data System Atlas 5th edition. Diagn Interv Radiol Ank Turk. Published Online First: 9 September 2024. 10.4274/dir.2024.242876

16. Jedrzejczak WW, Skarzynski PH, Raj-Koziak D, Sanfins MD, Hatzopoulos S, Kochanek K: ChatGPT for Tinnitus Information and Support: Response Accuracy and Retest after Three and Six Months. Brain Sci. 2024, 14:465. 10.3390/brainsci14050465

17. Wang S, Mo C, Chen Y, Dai X, Wang H, Shen X: Exploring the Performance of ChatGPT-4 in the Taiwan Audiologist Qualification Examination: Preliminary Observational Study Highlighting the Potential of AI Chatbots in Hearing Care. JMIR Med Educ. 2024, 10:e55595. 10.2196/55595

18. OpenAI Platform. Accessed: October 8, 2024. https://platform.openai.com.

19. Cheat Sheet: Mastering Temperature and Top_p in ChatGPT API - API. OpenAI Dev. Forum. (2023). Accessed: October 8, 2024. https://community.openai.com/t/cheat-sheet-mastering-temperature-and-top-p-in-chatgpt-api/172683.

20. Davis J, Van Bulck L, Durieux BN, Lindvall C: The Temperature Feature of ChatGPT: Modifying Creativity for Clinical Research. JMIR Hum Factors. 2024, 11:e53559. 10.2196/53559

21. Benjamini Y, Hochberg Y: Controlling the False Discovery Rate: A Practical and Powerful Approach to Multiple Testing. J R Stat Soc Ser B Stat Methodol. 1995, 57:289–300. 10.1111/j.2517-6161.1995.tb02031.x

22. Pilka E, Kochanek K, Jedrzejczak WW, Saczek A, Skarzynski H, Niedzielski A: Comparison of tympanometry results for probe tones of 226 Hz and 1000 Hz in newborns. Int J Pediatr Otorhinolaryngol. 2021, 147:110804. 10.1016/j.ijporl.2021.110804

23. Hoffmann A, Deuster D, Rosslau K, Knief A, Am Zehnhoff-Dinnesen A, Schmidt C-M: Feasibility of 1000 Hz tympanometry in infants: tympanometric trace classification and choice of probe tone in relation to age. Int J Pediatr Otorhinolaryngol. 2013, 77:1198–203. 10.1016/j.ijporl.2013.05.001

24. Liu Y, Ju S, Wang J: Exploring the potential of ChatGPT in medical dialogue summarization: a study on consistency with human preferences. BMC Med Inform Decis Mak. 2024, 24:75. 10.1186/s12911-024-02481-8

25. Bansal G, Chamola V, Hussain A, Guizani M, Niyato D: Transforming Conversations with AI—A Comprehensive Study of ChatGPT. Cogn Comput. 2024, 16:2487–510. 10.1007/s12559-023-10236-2

26. de Winter JCF, Driessen T, Dodou D: The use of ChatGPT for personality research: Administering questionnaires using generated personas. Personal Individ Differ. 2024, 228:112729. 10.1016/j.paid.2024.112729

27. Jedrzejczak WW, Kobosko J: Do Chatbots Exhibit Personality Traits? A Comparison of ChatGPT and Gemini. 2024. 10.31234/osf.io/xh9my

28. Bender EM, Gebru T, McMillan-Major A, Shmitchell S: On the Dangers of Stochastic Parrots: Can Language Models Be Too Big? In: Proceedings of the 2021 ACM Conference on Fairness, Accountability, and Transparency. Association for Computing Machinery: New York, NY, USA; 2021. 610–23. 10.1145/3442188.3445922

29. Ji Z, Lee N, Frieske R, et al.: Survey of Hallucination in Natural Language Generation. ACM Comput Surv. 2023, 55:248:1-248:38. 10.1145/3571730

30. Frosolini A, Franz L, Benedetti S, et al.: Assessing the accuracy of ChatGPT references in head and neck and ENT disciplines. Eur Arch Otorhinolaryngol. 2023, 280:5129–33. 10.1007/s00405-023-08205-4

31. Jędrzejczak WW, Pastucha M, Skarżyński H, Kochanek K: Comparison of ChatGPT and Gemini as sources of references in otorhinolaryngology. 2024, 2024.08.12.24311896. 10.1101/2024.08.12.24311896

