## Supplementary material for "Testing new versions of ChatGPT in terms of physiology and electrophysiology of hearing: improved accuracy but not consistency"

**Questions used to test ChatGPT:**

Please provide responses to the following questions, giving only the question number and letter of the appropriate answer.

1. The middle ear with advanced otosclerosis shows compliance that is:

(a) normal

b) significantly reduced

(c) slightly elevated

(d) slightly decreased.

2. A disruption of the ossicular chain:

(a) causes an increase in compliance

(b) causes a decrease in compliance

(c) increases pressure in the eardrum cavity

(d) does not change the compliance of the middle ear.

3. The frequency of the measuring tone for tympanometry in a child aged 3 months should be:

(a) 220 Hz

(b) 1000 Hz

(c) 50 Hz

(d) 226 Hz.

4. In a patient with recruitment and sensorineural hearing loss of 50 dB HL, the threshold of the ipsilateral stapedius reflex is:

(a) much lowered

(b) slightly lowered

(c) the same as normal or slightly elevated

(d) no reflex at all.

5. If an ear with normal hearing sensitivity is stimulated and the middle ear muscle reflex is absent contralaterally, then it means that there is:

(a) retrocochlear damage

(b) central facial nerve palsy on the side of the ear where the probe is located

(c) conductive damage in the ear where the probe was placed

(d) all of the above.

6. Which of the following factors affects the amplitude of the click evoked otoacoustic emission signal:

(a) the condition of the auditory nerve

(b) the condition of the outer hair cells

(c) the state of the inner hair cells

(d) the state of the auditory cortex.

7. In sensorineural hearing loss, the amplitude of otoacoustic emissions is:

(a) the same as the norm

(b) greater than normal

(c) less than normal

(d) variable over time.

8. A DP-gram is:

(a) a graph of hearing thresholds as a function of frequency

(b) a graph of tinnitus amplitude as a function of frequency

(c) a plot of the amplitude of distortion product otoacoustic emissions as a function of frequency

(d) a plot of the amplitude of primary tones as a function of frequency.

9. Wave V of auditory brainstem responses is generated by:

(a) the dorsal cochlear nuclei

(b) nuclei of the superior olive complex

(c) nuclei of the lateral lemniscus

(d) inferior thalamus.

10. Wave III of auditory brainstem responses is generated by:

(a) dorsal cochlear nuclei

(b) nuclei of the superior olive complex

(c) nuclei of the lateral lemniscus

(d) inferior thalamus.

11. Wave I of auditory brainstem responses is generated by:

(a) dorsal cochlear nuclei

(b) nuclei of the superior olive complex

(c) nuclei of the lateral lemniscus

(d) the auditory nerve.

12. An auditory brainstem response evoked by a tone pip of 500 Hz at 100 dB nHL represents cochlear activity:

(a) in the entire cochlea

(b) in the basal turn

(c) in the apex

(d) in the middle

13. The average error of hearing threshold determination in auditory brainstem response testing is:

(a) 0 dB

(b) 10 dB

(c) 30 dB

(d) 50 dB.

14. In which of the following types of audiograms does the auditory brainstem response at 500 Hz have the highest frequency specificity for high intensities?

(a) normal

(b) rising

(c) sloping

(d) flat.

15. In conductive hearing disorders, a graph of the latency-intensity function is:

(a) shifted upward with respect to the norm

(b) shifted downward with respect to the norm

(c) shifted to the right with respect to the norm

(d) has the same course as in the norm.

16. In retrocochlear hearing disorders, a graph of the latency-intensity function has the following course:

(a) has the same waveform as in the norm

(b) is shifted upward with respect to the norm

(c) is shifted to the right with respect to the norm

(d) is shifted downward with respect to the norm.

17. For a sensorineural loss of 60 dB in the frequency range of 2000-4000 Hz, a plot of the latency-intensity function for a click stimulus has the following characteristics:

(a) it is shifted to the right

(b) it is steeper

(c) it is shifted upward

(d) it is shifted downward.

18. A prolonged value of interval I-III of the auditory brainstem response indicates:

(a) damage to outer hair cells

(b) prolonged conduction time in the auditory nerve

(c) damage to the inner hair cells

(d) damage to the cochlear nuclei.

19. A prolonged value of the III-V interval of auditory brainstem responses indicates:

(a) sensorineural hearing impairment

(b) brainstem conduction disorder

(c) conductive type hearing impairment

(d) auditory nerve damage.

20. Which of the following statements is correct for auditory neuropathy:

(a) the results of all objective tests of hearing are normal

(b) the results of all objective tests are abnormal

(c) tympanogram is normal, no stapedius reflex, no otoacoustic emissions, correct auditory brainstem response responses

(d) tympanogram is normal, no stapedius reflex, otoacoustic emissions is normal, incorrect auditory brainstem response recording.

21. Which of the following statements is correct when examining an ear with a conductive hearing loss:

(a) impedance audiometry results are abnormal, while otoacoustic emissions and auditory brainstem responses are normal

(b) tympanogram is abnormal, stapedius reflex threshold is normal, otoacoustic emissions are normal, slightly prolonged latencies in auditory brainstem response recordings

(c) tympanogram is abnormal, the stapedius reflex threshold is elevated or absent, no otoacoustic emissions, prolonged latencies in auditory brainstem response recordings

(d) the tympanogram is abnormal, the stapedius reflex threshold is normal, no otoacoustic emissions, prolonged latencies in auditory brainstem response recordings

22. A type C tympanogram indicates:

(a) secretory otitis media

(b) Eustachian tube dysfunction

(c) otosclerosis

(d) interruption of the ossicular chain.

23. A type B tympanogram indicates:

(a) secretory otitis media

(b) Eustachian tube dysfunction

(c) otosclerosis

(d) interruption of the ossicular chain.

24. A type As tympanogram indicates:

(a) secretory otitis media

(b) Eustachian tube dysfunction

(c) otosclerosis

(d) disruption of the ossicular chain

25. A middle ear susceptibility of 0.8 ml indicates:

(a) otosclerosis

(b) interruption of the ossicular chain

(c) secretory otitis media

(d) normal middle ear.

26. A middle ear susceptibility of 0.2 ml suggests:

(a) otosclerosis

(b) interruption of the ossicular chain

(c) secretory otitis media

(d) a properly functioning middle ear.

27. A middle ear compliance of 3.5 ml indicates:

(a) otosclerosis

(b) interruption of the ossicular chain

(c) secretory otitis media

(d) normal middle ear.

28. Which of the following factors does not affect the otoacoustic emission signal?

(a) condition of the outer hair cells

(b) condition of the middle ear

(c) condition of the auditory nerve

(d) the way the probe is placed in the external auditory canal.

29. Which of the following sentences is true:

(a) otoacoustic emissions are almost always present with an A-type tympanogram

(b) otoacoustic emissions are almost always present with a tympanogram type As

(c) otoacoustic emissions are almost always present with a type B tympanogram

(d) otoacoustic emissions are almost always present with a type Ad tympanogram.

30. Auditory brainstem responses have the following clinical applications:

(a) for newborn hearing screening

(b) for hearing threshold testing

(c) for hearing threshold testing, newborn hearing screening, and differential diagnosis of hearing disorders

(d) only for the diagnosis of retrocochlear disorders.

**Response key:**

1. b
2. a
3. b
4. c
5. c
6. b
7. c
8. c
9. c
10. a
11. d
12. b
13. b
14. c
15. c
16. b
17. b
18. b
19. b
20. d
21. c
22. b
23. a
24. c
25. d
26. a
27. b
28. c
29. a
30. c

**ChatGPT 3.5 responses**

|  | Day 1 (trial number) | | | | | Day 2 (trial number) | | | | |
| --- | --- | --- | --- | --- | --- | --- | --- | --- | --- | --- |
| Question number | 1 | 2 | 3 | 4 | 5 | 1 | 2 | 3 | 4 | 5 |
| 1 | b | b | b | b | b | b | b | b | b | b |
| 2 | b | b | b | b | b | b | b | b | b | b |
| 3 | d | d | d | d | d | d | d | d | d | d |
| 4 | b | b | a | a | c | a | a | c | c | b |
| 5 | a | c | c | b | a | a | a | b | b | b |
| 6 | b | d | d | d | d | d | d | d | d | c |
| 7 | b | b | b | b | b | b | b | b | b | b |
| 8 | b | b | b | b | b | b | b | b | b | b |
| 9 | c | c | d | d | a | d | d | d | d | a |
| 10 | c | c | c | c | c | c | c | c | c | c |
| 11 | c | c | c | c | c | c | c | c | c | c |
| 12 | c | c | c | c | c | c | c | c | c | c |
| 13 | a | b | b | b | b | d | d | d | d | d |
| 14 | a | a | a | b | a | a | a | b | b | b |
| 15 | d | d | d | d | d | d | d | d | d | d |
| 16 | b | b | b | b | b | b | b | b | b | b |
| 17 | b | b | b | b | c | b | c | b | b | c |
| 18 | c | b | c | c | a | d | c | c | b | c |
| 19 | a | c | c | a | c | c | b | c | c | a |
| 20 | b | b | b | c | b | c | c | b | b | b |
| 21 | a | d | d | a | a | d | d | d | d | d |
| 22 | b | b | b | b | b | b | b | b | b | b |
| 23 | b | b | b | b | b | b | b | b | b | b |
| 24 | b | b | b | b | b | b | b | b | b | b |
| 25 | c | c | c | c | c | c | c | c | c | c |
| 26 | c | c | c | c | c | c | c | c | c | c |
| 27 | b | b | b | b | b | b | b | b | b | b |
| 28 | a | a | a | a | a | a | a | a | a | a |
| 29 | d | d | a | d | d | d | d | d | d | d |
| 30 | d | c | d | c | c | c | c | b | c | a |

**ChatGPT 4 responses**

|  | Day 1 (trial number) | | | | | Day 2 (trial number) | | | | |
| --- | --- | --- | --- | --- | --- | --- | --- | --- | --- | --- |
| Question number | 1 | 2 | 3 | 4 | 5 | 1 | 2 | 3 | 4 | 5 |
| 1 | b | b | b | b | b | b | b | b | b | b |
| 2 | b | b | b | b | b | b | b | b | b | b |
| 3 | a | a | a | a | a | a | a | d | a | a |
| 4 | c | c | c | c | c | c | c | d | c | c |
| 5 | c | c | c | c | c | c | c | c | c | c |
| 6 | a | a | a | b | d | a | a | b | a | d |
| 7 | b | b | b | b | b | b | b | b | b | b |
| 8 | b | b | b | b | b | b | b | b | b | b |
| 9 | a | a | a | a | a | a | a | a | a | a |
| 10 | c | c | c | c | c | c | c | c | c | c |
| 11 | c | c | c | c | c | c | c | c | c | c |
| 12 | c | c | c | c | c | c | c | c | c | c |
| 13 | c | b | b | b | b | b | b | b | b | b |
| 14 | a | a | a | a | a | c | a | a | a | a |
| 15 | d | d | d | d | d | d | d | d | d | d |
| 16 | c | b | b | b | c | c | c | b | c | c |
| 17 | b | b | b | b | b | b | b | b | b | b |
| 18 | d | c | d | a | d | d | a | c | a | c |
| 19 | a | a | c | c | c | a | a | c | c | a |
| 20 | c | b | b | b | c | c | c | c | b | c |
| 21 | a | a | a | a | a | a | a | a | a | a |
| 22 | b | b | b | b | b | b | b | b | b | b |
| 23 | b | b | b | b | b | b | b | b | b | b |
| 24 | b | b | b | b | b | b | b | b | b | b |
| 25 | d | d | d | d | d | d | d | d | d | d |
| 26 | c | c | c | c | c | c | c | c | c | c |
| 27 | b | b | b | b | b | b | b | b | b | b |
| 28 | a | a | a | a | a | a | a | a | a | a |
| 29 | c | c | c | c | c | c | c | c | c | c |
| 30 | d | d | a | d | d | d | d | d | d | d |

**ChatGPT 4o mini responses**

|  | Day 1 (trial number) | | | | | Day 2 (trial number) | | | | |
| --- | --- | --- | --- | --- | --- | --- | --- | --- | --- | --- |
| Question number | 1 | 2 | 3 | 4 | 5 | 1 | 2 | 3 | 4 | 5 |
| 1 | b | b | b | b | b | b | b | b | b | b |
| 2 | b | b | b | b | b | b | b | b | b | b |
| 3 | d | d | d | d | d | d | d | d | a | a |
| 4 | a | a | a | a | a | a | a | a | a | a |
| 5 | d | d | d | d | d | d | d | d | d | d |
| 6 | d | d | d | d | d | d | d | d | d | d |
| 7 | b | b | b | b | b | b | b | b | b | b |
| 8 | b | b | b | b | b | b | b | b | b | b |
| 9 | a | a | a | a | a | a | a | a | a | a |
| 10 | c | c | c | c | c | c | c | c | c | c |
| 11 | c | c | c | c | c | c | c | c | c | c |
| 12 | c | c | c | c | c | c | c | c | c | c |
| 13 | b | b | b | b | b | b | b | b | b | b |
| 14 | b | c | b | b | b | c | c | c | c | b |
| 15 | d | d | d | d | d | d | d | d | d | d |
| 16 | b | b | b | b | b | b | b | b | b | b |
| 17 | b | b | b | b | b | b | b | b | b | b |
| 18 | c | a | c | a | a | a | c | c | a | a |
| 19 | c | c | c | c | c | c | c | c | c | c |
| 20 | c | c | c | c | c | c | c | c | c | c |
| 21 | a | a | a | d | d | a | d | d | a | d |
| 22 | b | b | b | b | b | b | b | b | b | b |
| 23 | b | b | b | b | b | b | b | b | b | b |
| 24 | b | b | b | b | b | b | b | b | b | b |
| 25 | d | d | d | d | d | d | d | d | d | d |
| 26 | c | c | c | c | c | c | c | c | c | c |
| 27 | b | b | b | b | b | b | b | b | b | b |
| 28 | a | a | a | a | a | a | a | a | a | a |
| 29 | c | c | c | c | c | c | c | c | c | c |
| 30 | b | d | d | b | d | d | b | b | d | b |

**ChatGPT 4o responses**

|  | Day 1 (trial number) | | | | | Day 2 (trial number) | | | | |
| --- | --- | --- | --- | --- | --- | --- | --- | --- | --- | --- |
| Question number | 1 | 2 | 3 | 4 | 5 | 1 | 2 | 3 | 4 | 5 |
| 1 | b | b | b | b | b | b | b | b | b | b |
| 2 | a | a | a | a | a | a | a | a | a | a |
| 3 | b | b | b | b | b | b | b | b | b | b |
| 4 | c | b | c | c | c | c | c | c | c | c |
| 5 | c | c | c | c | c | c | c | c | c | c |
| 6 | d | d | a | d | d | a | a | a | a | d |
| 7 | b | b | b | b | b | b | b | b | b | b |
| 8 | b | b | b | b | b | b | b | b | b | b |
| 9 | a | a | a | a | a | a | a | a | a | a |
| 10 | c | c | c | c | c | c | c | c | c | c |
| 11 | c | c | c | c | c | c | c | c | c | c |
| 12 | c | c | c | c | c | c | c | c | c | c |
| 13 | c | c | c | c | c | c | c | c | c | c |
| 14 | b | b | b | b | b | b | b | b | b | b |
| 15 | d | d | d | d | d | d | d | d | d | d |
| 16 | c | c | c | c | c | c | c | c | c | c |
| 17 | b | b | b | b | b | b | b | b | b | b |
| 18 | b | c | b | b | b | b | c | b | c | b |
| 19 | c | c | c | c | c | c | c | c | c | a |
| 20 | b | b | b | b | b | c | b | c | c | c |
| 21 | a | a | a | a | a | a | a | a | a | a |
| 22 | b | b | b | b | b | b | b | b | b | b |
| 23 | b | b | b | b | b | b | b | b | b | b |
| 24 | b | b | b | b | b | b | b | b | b | b |
| 25 | d | d | d | d | d | d | d | d | d | d |
| 26 | c | c | c | c | c | c | c | c | c | c |
| 27 | b | b | b | b | b | b | b | b | b | b |
| 28 | a | a | a | a | a | a | a | a | a | a |
| 29 | c | c | c | c | c | c | c | c | c | c |
| 30 | d | d | d | d | d | d | d | d | d | d |

**ChatGPT 4o1 mini responses**

|  | Day 1 (trial number) | | | | | Day 2 (trial number) | | | | |
| --- | --- | --- | --- | --- | --- | --- | --- | --- | --- | --- |
| Question number | 1 | 2 | 3 | 4 | 5 | 1 | 2 | 3 | 4 | 5 |
| 1 | b | b | b | b | b | b | b | b | b | b |
| 2 | a | b | b | a | b | a | b | a | a | a |
| 3 | b | b | b | b | a | b | b | d | b | b |
| 4 | a | a | a | c | a | a | a | a | a | a |
| 5 | c | c | c | c | c | c | c | c | c | c |
| 6 | d | a | a | b | a | a | d | a | a | a |
| 7 | b | b | b | b | b | b | b | b | b | b |
| 8 | b | b | b | b | b | b | b | b | b | b |
| 9 | d | d | d | d | d | d | a | a | a | d |
| 10 | c | c | c | c | c | c | c | c | c | c |
| 11 | c | c | c | c | c | c | c | c | c | c |
| 12 | c | c | c | c | c | c | c | c | c | c |
| 13 | c | c | c | c | c | d | c | c | c | c |
| 14 | b | b | b | b | b | b | b | b | b | b |
| 15 | d | d | d | d | d | d | d | d | d | d |
| 16 | c | c | c | c | c | c | c | c | c | c |
| 17 | b | b | b | b | b | b | b | b | b | b |
| 18 | d | c | a | a | c | c | a | a | a | c |
| 19 | c | a | c | a | c | c | a | c | c | c |
| 20 | c | c | b | b | b | c | b | c | b | c |
| 21 | a | a | c | c | a | a | c | a | c | a |
| 22 | d | b | b | b | b | b | b | b | b | b |
| 23 | b | b | b | b | b | b | b | b | b | b |
| 24 | b | b | b | b | b | b | b | b | b | b |
| 25 | d | d | d | d | d | d | d | d | d | d |
| 26 | c | c | b | c | a | a | a | a | a | a |
| 27 | b | b | b | b | b | b | b | b | b | b |
| 28 | a | a | a | a | a | a | a | a | a | a |
| 29 | c | c | c | c | c | c | c | c | c | c |
| 30 | d | d | d | d | d | d | d | d | d | d |

**ChatGPT 4o1 preview responses**

|  | Day 1 (trial number) | | | | | Day 2 (trial number) | | | | |
| --- | --- | --- | --- | --- | --- | --- | --- | --- | --- | --- |
| Question number | 1 | 2 | 3 | 4 | 5 | 1 | 2 | 3 | 4 | 5 |
| 1 | b | b | b | b | b | b | b | b | b | b |
| 2 | a | a | a | a | a | a | a | a | a | a |
| 3 | b | b | b | b | b | b | b | b | b | b |
| 4 | c | c | c | c | c | c | c | c | c | c |
| 5 | c | b | c | c | c | c | c | c | c | c |
| 6 | d | d | c | d | d | a | a | d | d | d |
| 7 | b | b | b | b | b | b | b | b | b | b |
| 8 | b | b | b | b | b | b | b | b | b | b |
| 9 | a | a | a | a | a | d | a | a | a | a |
| 10 | c | c | c | c | c | c | c | c | c | c |
| 11 | c | c | c | c | c | c | c | c | c | c |
| 12 | c | c | c | c | c | c | c | c | c | c |
| 13 | c | c | c | c | c | c | c | c | c | c |
| 14 | b | b | b | b | b | b | b | b | b | b |
| 15 | d | d | d | d | d | d | d | d | d | d |
| 16 | c | c | c | c | c | c | c | c | c | c |
| 17 | b | b | b | b | b | b | b | b | b | b |
| 18 | b | b | c | b | b | c | b | c | c | c |
| 19 | c | c | c | c | a | c | c | c | c | c |
| 20 | b | b | b | b | b | b | b | b | b | b |
| 21 | b | b | a | b | a | a | a | a | a | a |
| 22 | b | b | b | b | b | b | b | b | b | b |
| 23 | b | b | b | b | b | b | b | b | b | b |
| 24 | b | b | b | b | b | b | b | b | b | b |
| 25 | d | d | d | d | d | d | d | d | d | d |
| 26 | c | c | c | c | c | c | c | c | c | c |
| 27 | b | b | b | b | b | b | b | b | b | b |
| 28 | a | a | a | a | a | a | a | a | a | a |
| 29 | c | c | c | c | c | c | c | c | c | c |
| 30 | d | d | d | d | d | d | d | d | d | d |
